# Imaging of increased peritumoral glutamate and glutamine in gliomas using 7T MRSI

**DOI:** 10.1101/2024.10.25.24316010

**Authors:** Gilbert Hangel, Philipp Lazen, Cornelius Cadrien, Stefanie Chambers, Julia Furtner, Lukas Hingerl, Bernhard Strasser, Barbara Kiesel, Mario Mischkulnig, Matthias Preusser, Thomas Roetzer-Pejrimovsky, Adelheid Wöhrer, Wolfgang Bogner, Karl Rössler, Siegfried Trattnig, Georg Widhalm

## Abstract

**Objectives:** Diffuse gliomas, due to their infiltrative properties, still lack effective treatment options. Recent research indicates that infiltration, malignancy, and symptoms such as epilepsy are related to synaptic connections between infiltrating glioma cells and cytotoxic levels of glutamate release. We previously showed that high-resolution 7T magnetic resonance spectroscopic imaging (MRSI) can resolve metabolic heterogeneities in gliomas. With this study, we evaluated 7T MRSI-derived glutamate (Gln) and glutamine (Glu) ratio maps for their use in defining infiltrative tumor activity in the peritumoral region.

**Materials and Methods:** We analyzed 7T MRSI scans of 36 patients with low- and high-grade gliomas. Within the visible tumor and a peritumoral shell, we calculated medians and Dice similarity coefficients (DSC) for nine metabolic ratios with and without hotspot thresholding and evaluated their correlation to and statistical significance between clinical parameters (e.g., tumor-associated epilepsy, IDH status, grade).

**Results:** The Glu/tCr (total creatine) median was significantly higher in the peritumoral VOI (1.13) compared to the tumor (0.92) and normal-appearing white matter (NAWM, 0.87), while the Gln/tCr median was highest in the tumor (0.77, vs 0.44 peritumoral and 0.33 in NAWM, all significantly different). Glu/tCho (total choline) was significantly higher in the peritumor as well (3.44 vs 2.23 tumoral and 2.06 in NAWM). Peritumoral DSCs for Glu/tCr and Gln/tCr hotspots were comparable (0.53 to 0.51). Peritumoral Gln/Glu was significantly different between patients with and without tumor-associated epilepsy, and intratumoral (Glu+Gln)/tCr was significantly different between IDH mutation and wildtype. IDH mutation correlated negatively with the intratumoral (Glu+Gln)/tCr median (-0.53) and high grade correlated with intratumoral Glx/tNAA, Glx/tCr, and Gln/tCr medians (0.50/0.53/0.58).

**Conclusions:** 7T MRSI can not only map relevant metabolic information in the structurally visible tumor volume, but also detect infiltration in the peritumoral area. Gln and Glu are candidates for the development of presurgical imaging and treatment monitoring.

## Introduction

Diffuse gliomas, brain tumors that originate from glial cells, make up the majority of malignant brain tumors^1^. Their infiltrative growth into the peritumoral brain parenchyma limits treatment options, such as resection and radiation therapy and causes high morbidity, e.g., epilepsy^2^, and mortality^3^. In clinical practice, structural MRI is used for the presurgical definition of the infiltration zone to allow a maximum safe resection. But infiltrating glioma cells remain beyond the MRI-visible changes, leading to recurrence.

As our understanding of molecular and metabolic processes of gliomas has grown, classification of gliomas relies increasingly on molecular markers. The 2021 WHO classification^4^ recognized classes of gliomas as “astrocytoma, IDH-mutant,” “oligodendroglioma, IDH-mutant, and 1p/19q-codeleted,” or “glioblastoma, IDH-wildtype.” Isocitrate dehydrogenase-mutations (IDH) lead to the production of the oncometabolite 2-hydroxyglutarate (2HG), whose role is a focus of research. Yet, 2HG is not the only molecule related to the citric acid cycle relevant to gliomas.

Glutamate (Glu) is a major excitatory neurotransmitter, but infiltrating glioma cells release amounts that are toxic to surrounding cells^5^, including causing necrosis^6^. Glioblastoma cells even move within peritumoral tissue^5^ and form synaptic connections to neurons that drive tumor growth^7,8^. This glutamate activity^9^ seems linked to AMPA receptors and calcium signaling in tumor and brain cells alike^5^. Similar changes in glutamate/calcium homeostasis were also reported for astrocytomas^10^. α- ketoglutarate (αKG) is the substrate for 2HG synthesis in IDH mutation, and can be converted from glutamate^11^. But in IDH wildtype tumors, conversely, αKG-sourced glutamate can be exchanged for extracellular cysteine (Cys), on the one hand contributing to the aforementioned toxic glutamate levels, and on the other hand boosting the synthesis of the antioxidant glutathione (GSH)^11^.

Glutamine (Gln) is formed from Glu and ammonia in astrocytes and acts as precursor of Glu and γ- amino butyric acid (GABA) in neurons^12^. Tumor cells show an increased dependence on Gln for metabolism, biosynthesis and signaling^13^. Gln, as Glu-precursor, is also a precursor of αKG^14^ and therefore 2HG^11^. Direct deamidation of Gln by glioma cells does not only release cytotoxic Glu, but also ammonia, which causes swelling/edema and inhibits the astrocytic glutamate uptake that delays glioma growth^15^. Glioblastoma Gln levels are higher in males than in females^16^.

One additional aspect is glioma epileptogenicity. Seizures are the most common symptom of gliomas at diagnosis^2^. The release of Glu by infiltrating glioma cells is fundamental to tumor-associated epilepsy (TAE)^17^. In human tumoral and peritumoral tissue samples, Glu concentrations and Glu transporter expression positively correlated with preoperative seizures^18^.

Based on these neurochemical findings for the role of Glu and Gln in gliomas, it would appear promising to image their concentrations *in vivo* both as a diagnostic measure and for research purposes. While both molecules can be detected with magnetic resonance spectroscopy (MRS), their spectral proximity makes them difficult to separate, necessitating to report the sum of Glu+Gln as Glx^19^. The general limitations of MRS with regard to SNR, scan times and resolution add to this^20^. Nonetheless, limited research on Glu and Gln in gliomas has been published. For 3T magnetic resonance spectroscopic imaging (MRSI), reduced ratios of Glu to total creatine (Glu/tCr) and increased Gln/tCr compared to normal-appearing white matter (NAWM) have been reported^21^. Intratumoral Glx concentrations detected with 3 Tesla (3T) single voxel MRS (SVS) were found to be significantly different between grades 2, 3, and 4 gliomas in the 2017 WHO classification^19^ and higher Glu/tCr significantly correlated with shorter progression-free survival and overall survival^22^. More 3T SVS glioblastoma studies found that an intratumoral Glu/tCr cutoff of >1.81 predicted postsurgical seizures with an area under the curve of 0.82^23^ and that peritumoral Glu/tCr was significantly increased in patients with TAE^24^. In sum, peritumoral Glu/Gln remains sparsely researched due to the resolution/coverage limitations of standard 3T MRS methods.

A new generation of MRSI that combines enhanced SNR and spectral separation at 7 Tesla (7T) with fast spatial-spectral encoding and free induction decay (FID) acquisition^25^ allows the acquisition of 3D neurochemical maps withing 15 minutes at 3.4mm isotropic resolution and has found intratumoral heterogeneities for Glu, Gln, total choline (tCho), and others such as glycine^26,27^. As this MRSI method can map not only Gln and Glu separately, but also image tumor and peritumor within one scan, it can investigate Glu/Gln changes as described by glioma research. Such detectable changes could be the basis for further research on the role of Glu in glioma infiltration and epileptogenesis.

### Purpose

Our purpose was to reinvestigate our previous high-resolution 3D MRSI scans of glioma patients with a focus on glutamate and glutamine, and to identify significant differences between tumor and peritumor, as well as the correlation of Glu and Gln to TAE and other tumor properties.

## Materials and Methods

### Subjects and clinical data

We used the datasets of 36 glioma patients (mean age 52 years, range 26-77 years, 14 female, Table 1) who had previously participated in 7T MRSI scans^28^ with IRB approval and written, informed consent. The cohort consisted of 16 glioblastoma, IDH-wildtype; 14 astrocytoma, IDH-mutant; and six oligodendroglioma, IDH-mutant, and 1p/19q-codeleted / 28 high-grade (i.e., grade 3 or 4) and eight low-grade, i.e., grade 2, gliomas. For the 7T scans, inclusion criteria were a Karnofsky performance status ≥ 70 and exclusion criteria were 7T MRI contraindications (e.g., pregnancy, claustrophobia, ferromagnetic implants, non-ferromagnetic metal head implants >12 mm). For the inclusion into this study, additional criteria were histologically confirmed gliomas (2021 WHO classification^29^, Table 1) and sufficient 7T data quality, as detailed in Cadrien et al. 2024^28^.

**Table 1:** An overview of the included patients and their evaluated molecular-pathological tumor properties according to the WHO 2021 classification. 1p/19q-codeletion = combined loss of alleles on chromosomes 1 and 19; CDKN2A/B = cyclin-dependent kinase inhibitor 2A/B; IDH = isocitrate dehydrogenase; IQR = inter-quartile range; TERT = telomerase reverse transcriptase [with C228T, C250T as possible TERT mutations]; WT = wild type.

| # | WHO 2021 Classification | Grade | IDH | 1p/19q-codeletion | TERT promoter mutation | CDKN2A/B homozygous deletion | age range | sex |
| --- | --- | --- | --- | --- | --- | --- | --- | --- |
| 1 | Astrocytoma, IDH-mutant | 2 | IDH1-mut | no | no | no | 31-40 | male |
| 2 | Astrocytoma, IDH-mutant | 4 | IDH1-mut | no | C250T | no | 51-60 | male |
| 3 | Astrocytoma, IDH-mutant | 4 | IDH1-mut | no | C250T | no | 51-60 | male |
| 4 | Glioblastoma, IDH-wildtype | 4 | WT | N/A | N/A | N/A | 41-50 | female |
| 5 | Astrocytoma, IDH-mutant | 3 | IDH1-mut | no | no | no | 41-50 | female |
| 6 | Astrocytoma, IDH-mutant | 3 | IDH1-mut | no | no | no | 21-30 | male |
| 7 | Glioblastoma, IDH-wildtype | 4 | WT | N/A | no | N/A | 51-60 | male |
| 8 | Astrocytoma, IDH-mutant | 2 | IDH1-mut | no | no | no | 31-40 | male |
| 9 | Glioblastoma, IDH-wildtype | 4 | WT | no | no | no | 51-60 | male |
| 10 | Astrocytoma, IDH-mutant | 2 | IDH1-mut | no | no | no | 71-80 | female |
| 11 | Glioblastoma, IDH-wildtype | 4 | WT | no | no | no | 31-40 | female |
| 12 | Glioblastoma, IDH-wildtype | 4 | WT | N/A | C228T | N/A | 71-80 | male |
| 13 | Oligodendroglioma, IDH-mutant, and 1p/19q-codeleted | 3 | IDH1-mut | yes | C228T | no | 51-60 | male |
| 14 | Astrocytoma, IDH-mutant | 3 | IDH1-mut | no | no | no | 61-70 | male |
| 15 | Glioblastoma, IDH-wildtype | 4 | WT | no | no | yes | 21-30 | female |
| 16 | Astrocytoma, IDH-mutant | 2 | IDH1-mut | no | no | no | 31-40 | male |
| 17 | Oligodendroglioma, IDH-mutant, and 1p/19q-codeleted | 3 | IDH1-mut | yes | N/A | yes | 51-60 | male |
| 18 | Glioblastoma, IDH-wildtype | 4 | WT | N/A | N/A | N/A | 61-70 | male |
| 19 | Glioblastoma, IDH-wildtype | 4 | WT | no | C250T | N/A | 51-60 | male |
| 20 | Astrocytoma, IDH-mutant | 3 | IDH1-mut | no | no | no | 21-30 | female |
| 21 | Glioblastoma, IDH-wildtype | 4 | WT | N/A | N/A | N/A | 61-70 | male |
| 22 | Oligodendroglioma, IDH-mutant, and 1p/19q-codeleted | 2 | IDH1-mut | yes | N/A | N/A | 41-50 | female |
| 23 | Oligodendroglioma, IDH-mutant, and 1p/19q-codeleted | 2 | IDH1-mut | yes | no | no | 31-40 | female |
| 24 | Oligodendroglioma, IDH-mutant, and 1p/19q-codeleted | 2 | IDH1-mut | yes | C250T | no | 61-70 | male |
| 25 | Glioblastoma, IDH-wildtype | 4 | WT | N/A | N/A | N/A | 71-80 | female |
| 26 | Astrocytoma, IDH-mutant | 2 | IDH1-mut | no | no | no | 31-40 | male |
| 27 | Glioblastoma, IDH-wildtype | 4 | WT | N/A | N/A | N/A | 71-80 | female |
| 28 | Glioblastoma, IDH-wildtype | 4 | WT | N/A | N/A | N/A | 51-60 | male |
| 29 | Oligodendroglioma, IDH-mutant, and 1p/19q-codeleted | 3 | IDH1-mut | yes | N/A | N/A | 51-60 | female |
| 30 | Astrocytoma, IDH-mutant | 3 | IDH1-mut | no | no | no | 31-40 | male |
| 31 | Glioblastoma, IDH-wildtype | 4 | WT | N/A | N/A | N/A | 51-60 | male |
| 32 | Astrocytoma, IDH-mutant | 4 | IDH1-mut | no | no | N/A | 21-30 | female |
| 33 | Glioblastoma, IDH-wildtype | 4 | WT | no | C228T | yes | 71-80 | male |
| 34 | Astrocytoma, IDH-mutant | 4 | IDH1-mut | no | no | N/A | 21-30 | female |
| 35 | Glioblastoma, IDH-wildtype | 4 | WT | N/A | N/A | N/A | 51-60 | male |
| 36 | Glioblastoma, IDH-wildtype | 4 | WT | no | no | N/A | 41-50 | female |

52±23 Median+IQR

**Table 2:** Median values (1st and 3rd quartile in brackets) of all evaluated ratios over the **TU1.5**, **PT1.5**, **TU0**, **PT0**, and **NAWM** VOIs. Of note, Glu/tCr and Glu/tCho are higher in **PT0** than in **TU0** and **NAWM**. Glu = glutamate; Gln = glutamine; Glx = glutamate + glutamine; NAWM = normal-appearing white matter; PT0 = peritumor segmentation without ratio threshold; PT1.5 = peritumor segmentation with 1.5×NAWM threshold; tCho = total choline; tCr = total creatine; tNAA = total N-acetyl aspartate; TU0 = tumor segmentation without ratio threshold; TU1.5 = tumor segmentation with 1.5×NAWM threshold; I = volume of interest.

| Metabolite Median | TU1.5 | PT1.5 | TU0 | PT0 | NAWM |
| --- | --- | --- | --- | --- | --- |
| **Glu/tNAA** | 0.98 [1.08-0.83] | 0.88 [0.98-0.80] | 0.72 [0.79-0.54] | 0.61 [0.68-0.55] | 0.42 [0.47-0.37] |
| **Gln/tNAA** | 0.70 [0.84-0.56] | 0.40 [0.54-0.36] | 0.61 [0.73-0.45] | 0.26 [0.29-0.22] | 0.18 [0.20-0.12] |
| **Glu/tCr** | 1.68 [1.93-1.52] | 1.67 [1.98-1.51] | 0.92 [1.08-0.83] | 1.13 [1.25-0.97] | 0.87 [1.01-0.74] |
| **Gln/tCr** | 0.99 [1.26-0.78] | 0.78 [1.01-0.67] | 0.77 [0.99-0.58] | 0.44 [0.51-0.39] | 0.33 [0.43-0.27] |
| **Glx/tNAA** | 1.46 [1.73-1.28] | 1.18 [1.31-1.14] | 1.26 [1.47-1.03] | 0.85 [0.93-0.76] | 0.56 [0.63-0.50] |
| **Glx/tCr** | 2.49 [2.77-2.07] | 2.38 [2.80-2.04] | 1.60 [1.98-1.35] | 1.56 [1.69-1.40] | 1.20 [1.38-1.01] |
| **Glu/tCho** | 4.34 [4.96-3.80] | 4.51 [5.49-4.21] | 2.23 [3.09-1.78] | 3.44 [3.94-2.83] | 2.06 [2.48-1.81] |
| **Gln/Glu** | 1.18 [1.27-0.98] | 0.86 [1.09-0.75] | 0.81 [1.01-0.61] | 0.41 [0.46-0.35] | 0.39 [0.49-0.32] |
| **tCho/tNAA** | 0.49 [0.55-0.41] | 0.39 [0.44-0.33] | 0.29 [0.41-0.21] | 0.18 [0.20-0.16] | 0.20 [0.22-0.17] |

**Table 3:** Distribution of p-values for median ratios between different VOIs. All thresholds were modified by a Bonferroni correction of 9 to account for multiple testing. Green highlights p<0.00011 (0.001 before correction), pale green indicates p<0.0055 (0.05 before correction), yellow means non- significant after correction, and red shows non-significance even before correction. In sum, these results show that Glu and Gln ratios in most cases can discern between tumor, peritumor, and NAWM. Gln/tNAA and Glx/tNAA do so in all cases while the reference tCho/tNAA cannot discern between NAWM and peritumor. Glu = glutamate; Gln = glutamine; Glx = glutamate + glutamine; NAWM = normal-appearing white matter; PT0 = peritumor segmentation without ratio threshold; PT1.5 = peritumor segmentation with 1.5×NAWM threshold; tCho = total choline; tCr = total creatine; tNAA = total N-acetyl aspartate; TU0 = tumor segmentation without ratio threshold; TU1.5 = tumor segmentation with 1.5×NAWM threshold; VOI = volume of interest.

| p-values of compared VOIs | Glu/tNAA | Gln/tNAA | Glu/tCr | Gln/tCr | Glx/tNAA | Glx/tCr | Glu/tCho | Gln/Glu | Cho/tNAA |
| --- | --- | --- | --- | --- | --- | --- | --- | --- | --- |
| **TU1.5 to PT1.5** | 0.00014457 | 2.85522E-09 | 0.72772871 | 0.00585915 | 1.01511E-07 | 0.58278507 | 6.6088E-06 | 9.4039E-06 | 1.1876E-05 |
| **TU0 to PT0** | 0.00543169 | 2.01992E-10 | 2.4044E-05 | 1.0096E-09 | 1.12678E-08 | 0.02158697 | 9.3276E-11 | 2.7319E-10 | 4.0299E-06 |
| **TU0 to NAWM** | 2.0395E-10 | 9.96081E-12 | 0.06918513 | 3.0237E-10 | 1.20888E-12 | 4.2003E-07 | 0.44881421 | 4.228E-09 | 3.0259E-05 |
| **PT0 to NAWM** | 2.2453E-13 | 3.59857E-07 | 1.135E-07 | 0.00032492 | 8.26724E-14 | 3.0889E-08 | 5.7076E-11 | 0.82019608 | 0.56111151 |

significant after Bonferroni-correction with $p < 0.00011$
significant after Bonferroni-correction with $p < 0.0055$
not significant after Bonferroni-correction
not significant before Bonferroni-correction

**Table 4:**
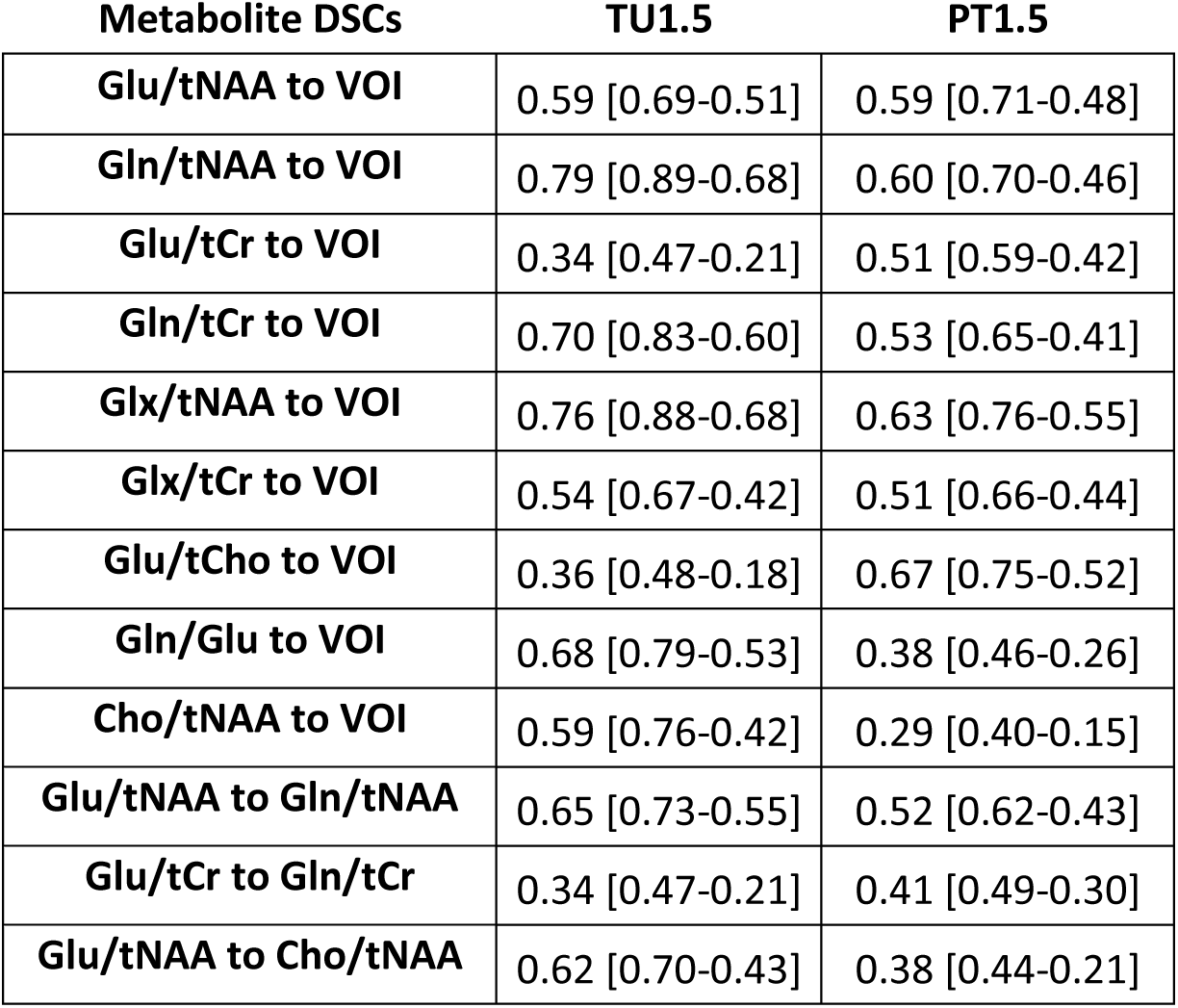
Median DSCs (1st and 3rd quartile in brackets) for the **TU1.5** and **PT1.5** thresholds. Gln/tNAA to the tumor segmentation VOI scores highest, while the reference ratio tCho/tNAA to the peritumoral segmentation scores lowest. DSC = Dice similarity coefficient; Glu = glutamate; Gln = glutamine; Glx = glutamate + glutamine; NAWM = normal-appearing white matter; PT1.5 = peritumor segmentation with 1.5×NAWM threshold; tCho = total choline; tCr = total creatine; tNAA = total N-acetyl aspartate; TU1.5 = tumor segmentation with 1.5×NAWM threshold; VOI = volume of interest.

| Metabolite DSCs | TU1.5 | PT1.5 |
| --- | --- | --- |
| **Glu/tNAA to VOI** | 0.59 [0.69-0.51] | 0.59 [0.71-0.48] |
| **Gln/tNAA to VOI** | 0.79 [0.89-0.68] | 0.60 [0.70-0.46] |
| **Glu/tCr to VOI** | 0.34 [0.47-0.21] | 0.51 [0.59-0.42] |
| **Gln/tCr to VOI** | 0.70 [0.83-0.60] | 0.53 [0.65-0.41] |
| **Glx/tNAA to VOI** | 0.76 [0.88-0.68] | 0.63 [0.76-0.55] |
| **Glx/tCr to VOI** | 0.54 [0.67-0.42] | 0.51 [0.66-0.44] |
| **Glu/tCho to VOI** | 0.36 [0.48-0.18] | 0.67 [0.75-0.52] |
| **Gln/Glu to VOI** | 0.68 [0.79-0.53] | 0.38 [0.46-0.26] |
| **Cho/tNAA to VOI** | 0.59 [0.76-0.42] | 0.29 [0.40-0.15] |
| **Glu/tNAA to Gln/tNAA** | 0.65 [0.73-0.55] | 0.52 [0.62-0.43] |
| **Glu/tCr to Gln/tCr** | 0.34 [0.47-0.21] | 0.41 [0.49-0.30] |
| **Glu/tNAA to Cho/tNAA** | 0.62 [0.70-0.43] | 0.38 [0.44-0.21] |

We further collected preoperative MRI data consisting of T1w with/without contrast-enhancement (Gadoteridol,0.1 mmol/kg), T2w, and T2w Fluid-attenuated inversion recovery MRI (FLAIR).

### 7T MRSI measurement and processing

We conducted all MRSI scans on a 7T scanner (Siemens Healthineers, Erlangen, Germany, Magnetom/Magnetom.plus after an upgrade) with a 1Tx/32Rx head coil (Nova Medical, Wilmington, MA, USA). Prior to MRSI, we acquired T1w-MRI (MP2RAGE) and FLAIR.

Our MRSI sequence^25,26,28^ used free-induction-decay acquisition and concentric ring trajectories to acquire a 64×64×39 matrix (FOV of 220×220×133mm³, resolution of 3.4×3.4×3.4mm³) in 15 minutes. Further parameters were TR 450 ms, acquisition delay 1.3 ms, 39° excitation flip angle, 345ms readout duration with 2778Hz spectral bandwidth, 7T-optimized WET water suppression^30^, and FOV placement maximizing tumor coverage in the superior parts of the brain which are less affected by B0- inhomogeneities.

We processed the MRSI raw data offline in a custom pipeline including iMUSICAL coil combination^25^, k-space re-gridding^25^ and coil-wise L2-regularization^31^ to remove lipid signals. The resulting spectra were quantified per voxel using LCModel (basis set: tCr, Cys, GABA, Gln, Glu, glycine, GSH, tCho, myo-inositol, serine, taurine, N-acetyl aspartate (NAA), NAA-glutamate, macromolecular baseline; 1.8-4.1 ppm range; separation performance of Glu and Gln were discussed previously^26^). The resulting intensities (in institutional units) were aggregated into 3D metabolic maps. As MRSI concentration estimation within the tumor was not possible without a knowledge of intratumoral voxel T1 and water concentrations, we calculated ratio maps. For our evaluation, we selected the ratios of Glu/ tNAA (NAA+NAAG), Gln/tNAA, Glu/tCr, Gln/tCr, Glx/tNAA, Glx/tCr, Glu/tCho, and Gln/Glu to investigate tumoral/peritumoral activity of Glu and Gln. As reference, we added the clinical standard ratio of Cho/tNAA.

For spectral quality evaluation, we calculated, voxel-wise, full-width-at-half-maximum (FHWM) and signal-to-noise ratio (SNR) of tCr at 3.02 ppm, as well as Cramér–Rao lower bounds (CRLB) of all metabolites^32^. More details, such as voxel exclusion criteria are summarized according to the MRSinMRS standard in Supplementary Table 1^33^ and described in previous publications^25,26,28^.

### Co-registration and VOI definition

We co-registered 3T and 7T images using MITK (Medical Imaging Interaction Toolkit) before a neuroradiologist (J.F.) segmented the tumor volumes (**TU**, binary “visible tumor including edema yes/no”) based on the 3T protocols^26^ in ITK-SNAP. We then resampled **TU** to the MRSI resolution. We then defined a peritumoral VOI (**PT**) made up of six iterations of dilation (corresponding to 2cm) of TU. Using 7T-T1w images for a gray matter (GM)/white matter (WM) segmentation, we defined a **NAWM** reference VOI by removing **TU** and **PT** from WM and eroding the result once. We also removed GM voxels from PT to allow a WM-centric comparison without higher-Glu GM effects.

For the subsequent evaluation using a python script, we used five VOIs per metabolite ratio: **NAWM** as above including ratios 0<ratiovoxel<10 (to avoid outliers); **TU0** and **PT0** using the same range to evaluate overall VOI trends; and **TU1.5** / **PT1.5**, which used 1.5*[median NAWM ratio per metabolite]< ratiovoxel<10 to look at metabolic hotspots.

### Analysis and evaluation

In the selected ratios, we conducted four analyses: 1) for statistical differences of median ratios between VOIs; 2) categorical differences per ratio and VOI; 3) Sørensen–Dice similarity coefficients (DSCs) between **TU1.5** and **PT1.5**; and 4) correlations between categories and ratios. We applied a Bonferroni-correction of 9 to all statistical test thresholds (0.05è0.0055; 0.001è0.00011).

### Differences between VOI medians

For all five VOIs and all nine ratios, we calculated median ratios. We applied paired two-sided t-tests for the median differences between the VOI pairs **TU1.5 – PT1.5**, **TU0 – PT0**, **TU0 – NAWM**, and **PT0 – NAWM**. We excluded other combinations, as they would be significantly different due to different thresholding.

### Categorical differences

For all four tumor VOIs and all selected ratios, we evaluated paired two-sided t-tests for the categorical pairs “TAE Yes/no,” “IDH mutation / IDH wildtype,” “Low grade / high grade,” “astrocytoma / oligodendroglioma,” “astrocytoma / glioblastoma,” and “oligodendroglioma / glioblastoma.”

### Similarity coefficients

We calculated patient-wise DSCs of the **TU1.5** and **PT1.5** VOIs from the original **TU** and **PT** segmentations, as well as the DSCs of Glu/tNAA to Gln/tNAA, Glu2tCr to Gln/tCr, and Glu/tNAA to Cho/tNAA using the following formula:

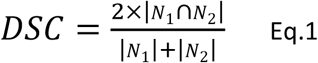

with |*N*_1_| and |*N*_2_| as voxel amounts. A DSC of 1 describes a complete overlap of two VOIs while a DSC of 0 would signify none. We then calculated median DSCs for the patient cohort.

### Correlations

Last, we computed correlation coefficients of the ratio and DSC medians to patient age, sex, grade (2,3,4), grade (low/high), IDH1 mutation status, TAE and peritumoral seizure onset. We did this for the VOIs **TU0**, **TU1.5**, **PT0**, and **PT1.5** with correlation coefficients <-0.5 or >0.5 defined as relevant findings.

## Results

In Fig.1, we present an example of Glu/tNAA, Gln/tNAA, and tCho/tNAA highlighting peritumoral regions in patient #4. Sup.Fig.1 shows example spectra for **TU**, **PT** and **NAWM**. Sup.Tbls 2-5 contain the patient-wise medians, DSCs, and thresholds used in this analysis.

**Figure 1:**
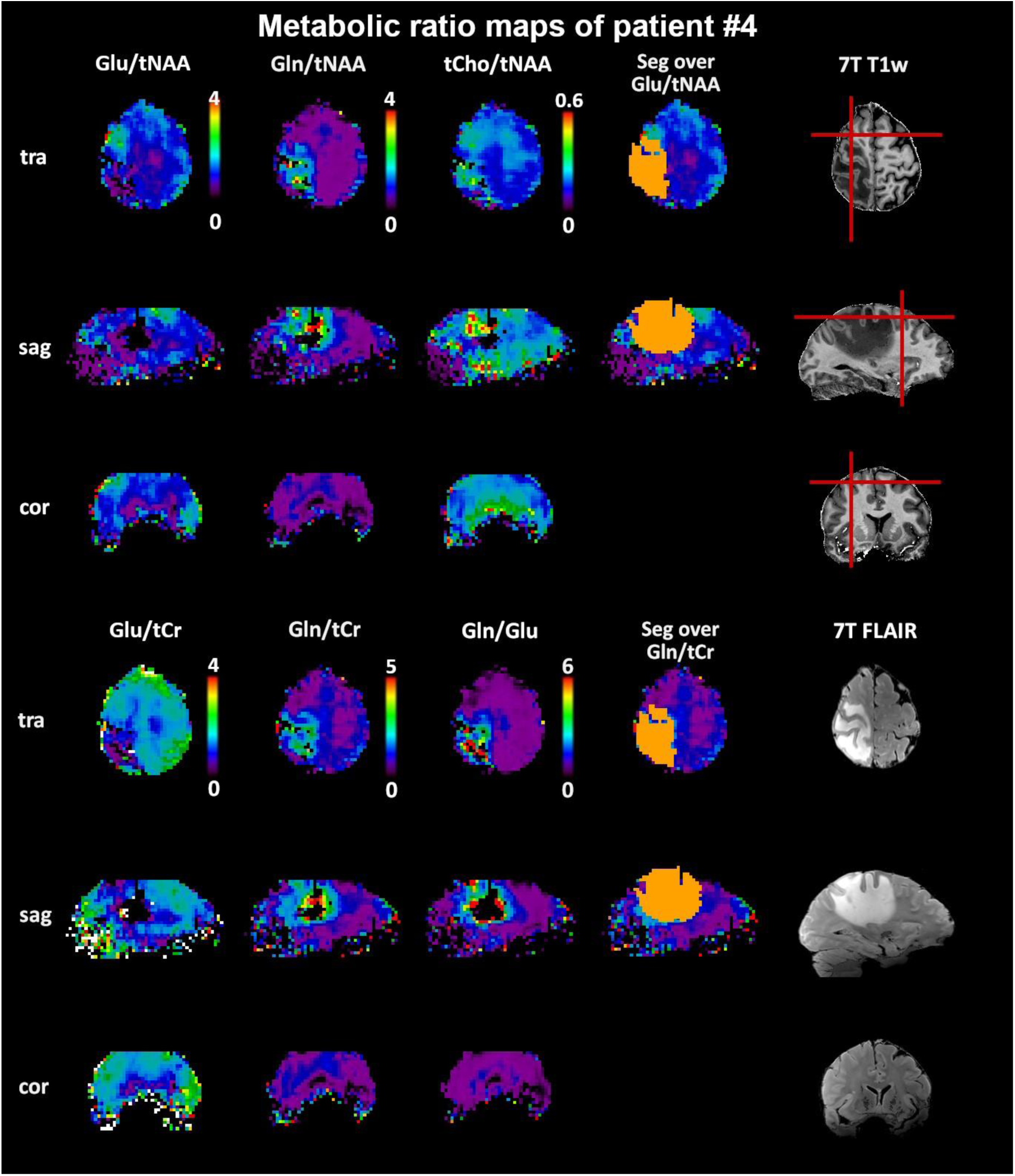
Exemplary metabolic ratio maps in patient #4 that display tumoral and peritumoral Glu and Gln distributions. Of note is a frontal peritumoral Glu/tNAA hotspot that has no correspondence on the Gln ratio maps. These maps also show the general power of Gln -based maps to discern tumor metabolism vs the healthy-appearing brain with a contrast of over a magnitude in this case. Red lines indicate the location of the MRSI slices. Cor = coronal; Glu = glutamate; Gln = glutamine; Seg = tumor segmentation; T1w = T1-weighted; sag = sagittal; tCho = total choline; tCr = total creatine; tra = transversal; tNAA = total N-acetyl aspartate.

### Differences between VOI medians

Median **TU** ratios were higher than **PT** ratios for both thresholds, except for Glu/tCr in **TU0** vs **PT0** and Glu/tCho for both (Tbl.2). All **TU/PT** VOIs were higher than in NAWM for the 0 threshold except tCho/tNAA in **PT0** vs **NAWM** (and obviously so for the 1.5 threshold). These distributions are also plotted in Fig.2, visualizing the differences between ratios to tCr and tNAA. The differences between Glu/tNAA and Glu/tCr show the impact of the chosen reference metabolite, as the tumoral decrease of tNAA masks the apparent increase of Glu in **PT0**. Combining Glu/tCr and Gln/tCr into Glx/tCr masks their diverging distributions. High similarity between the Gln/Glu and tCho/tNAA distributions was visible as well.

**Figure 2:**
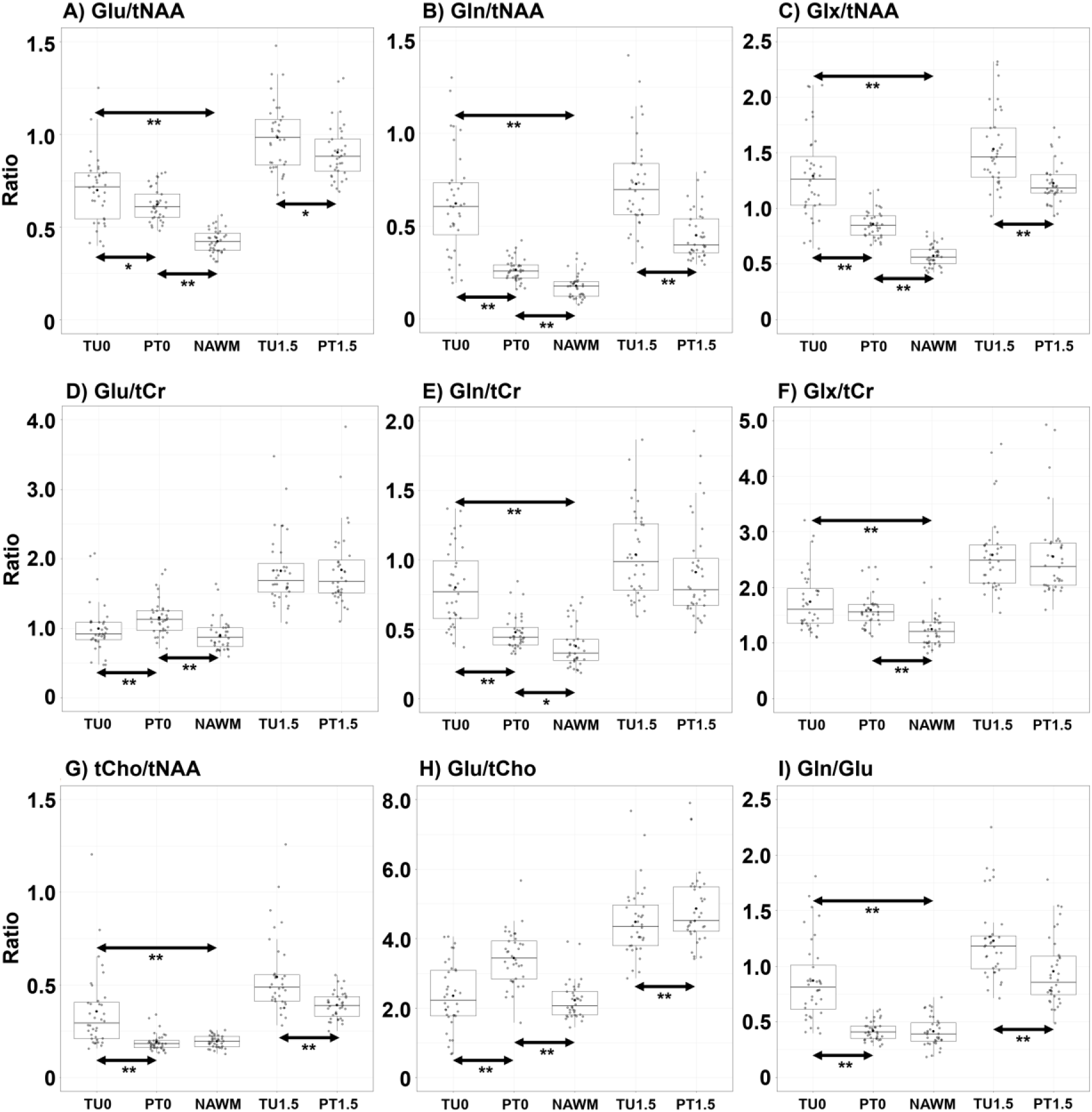
Box-plots of all evaluated ratios over the **TU1.5**, **PT1.5**, **TU0**, **PT0**, and **NAWM** VOIs that include statistically significant differences. * indicates p<0.0055 and ** indicates p<0.00011. All ratios could discern at least two compartments. Glu = glutamate; Gln = glutamine; Glx = glutamate + glutamine; NAWM = normal-appearing white matter; PT0 = peritumor segmentation without ratio threshold; PT1.5 = peritumor segmentation with 1.5×NAWM threshold; tCho = total choline; tCr = total creatine; tNAA = total N-acetyl aspartate; TU0 = tumor segmentation without ratio threshold; TU1.5 = tumor segmentation with 1.5×NAWM threshold; VOI = volume of interest.

The statistical significance of these differences (Tbl.3, Fig.2) elaborate these findings. The reference ratio could distinguish **TU0** from **PT0** and **NAWM**, but not between **PT0** and **NAWM**. All ratios with Glu and Gln except Gln/Glu significantly differed between **PT0** and **NAWM**, with Glx/tNAA having the lowest p-value and Glu/tCho the lowest for the non-tNAA ratios. For **TU0** and **NAWM**, all ratios except Glu/tCr and Glu/tCho were significantly different, with Gln/tNAA the most significant. All ratios, except

Glx/tCr, could significantly distinguish between **PT0** and **TU0**, with Glu/tCho the most significant. The thresholded **PT1.5** and **TU1.5** VOIs had Glu/tCr, Gln/tCr, and Glx/tCr as nonsignificant ratios and Gln/tCr as the most significantly different ratio. Overall, Gln ratios showed more significant differences than Glu ratios except for **PT0** to **NAWM** and tended to be more significant than tCho to tNAA.

### Categorical differences

We found significant differences for the categories “TAE Yes/no” in Gln/Glu **PT1.5** with p=0.0029, “IDH mutation / IDH wildtype” in Glx/tCr **TU0** with p=0.0009, and “astrocytoma / oligodendroglioma” in Gln/tNAA **PT0** with p=0.0023, as well as Glx/tNAA **PT0** with p=0.0023. These four distributions are plotted in Fig.3. Fig.4 gives an example of such differences for epilepsy and astrocytoma vs oligodendroglioma.

**Figure 3:**
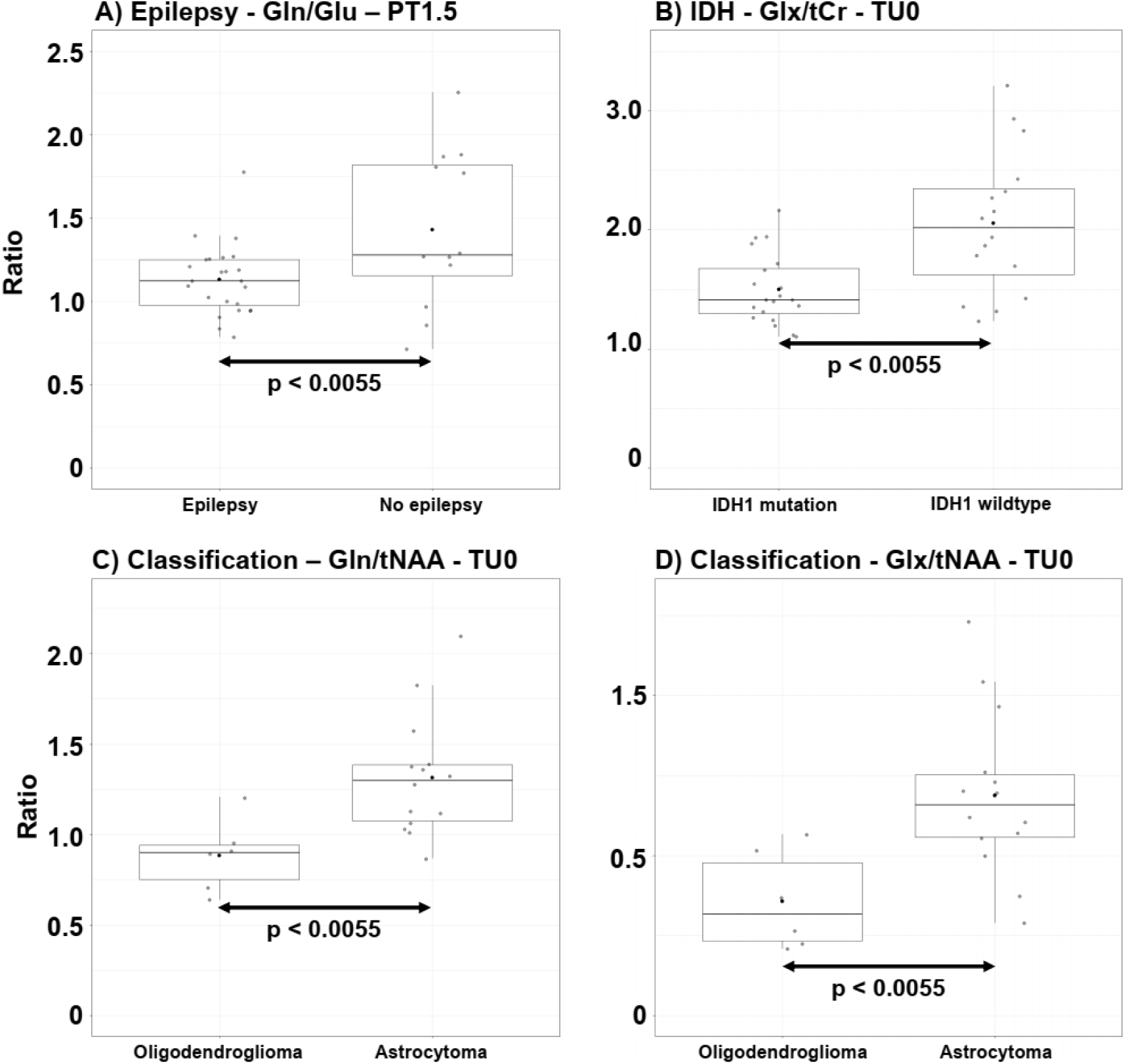
Box-plots of the four ratios for which we found significant categorical differences (tumor- associated epilepsy, IDH-mutation, classification). Glu = glutamate; Gln = glutamine; Glx = glutamate + glutamine; NAWM = normal-appearing white matter; PT1.5 = peritumor segmentation with 1.5×NAWM threshold; tCho = total choline; tCr = total creatine; tNAA = total N-acetyl aspartate; TU0 = tumor segmentation without ratio threshold.

**Figure 4:**
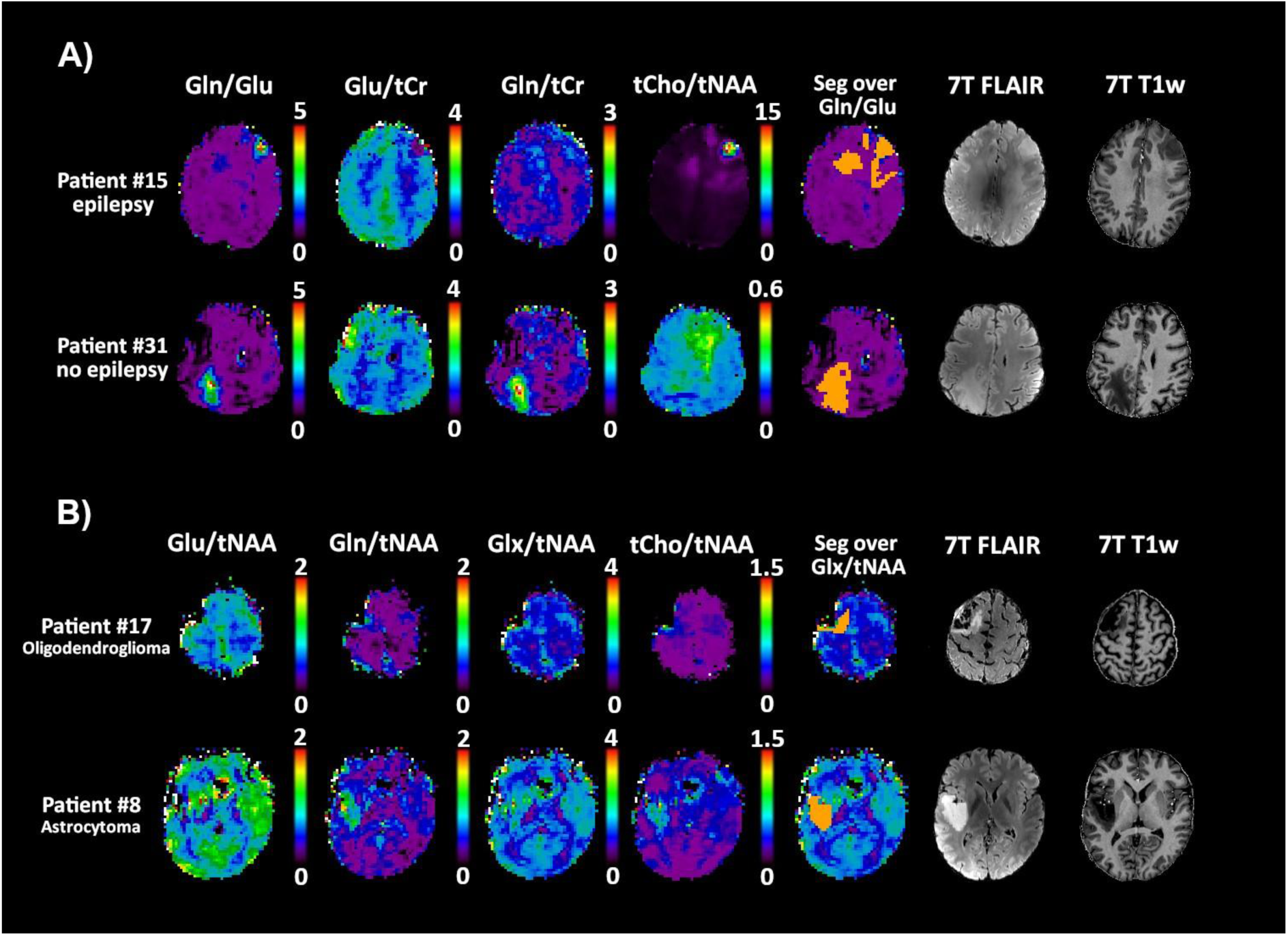
Comparison of transversal metabolic ratio maps between A) patients with and without epilepsy where median tumoral Gln/Glu ratios differed and B) oligodendroglioma, IDH-mutant, and 1p/19q-codeleted and astrocytoma, IDH-mutant, where median peritumoral Gln/tNAA and Glx/tNAA ratios differed. FLAIR = fluid attenuated inversion recovery; Glu = glutamate; Gln = glutamine; Glx = glutamate + glutamine; NAWM = normal-appearing white matter; PT1.5 = peritumor segmentation with 1.5×NAWM threshold; Seg = tumor segmentation; T1w = T1-weighted; tCho = total choline; tCr = total creatine; tNAA = total N-acetyl aspartate; TU1.5 = tumor segmentation with 1.5×NAWM threshold; VOI = volume of interest.

### Similarity coefficients

Of all resulting DSCs to the **TU** and **PT** segmentation VOIs (representing how much the thresholded hotspot filled them out, Tbl4., Fig.5), Gln/tNAA had the highest DSC even greater than tCho/tNAA, consistent with our previous research. DSCs to **TU** are generally higher than those to **PT**, except for Glu/tCr and Glu/tCho. Glu ratio DSCs are lower than Gln DSCs, but are generally higher in **PT** than in **TU**. For the DSCs between ratios, Glu/tNAA to Gln/tNAA was 0.65 in **TU1.5**, 0.52 in **PT1.5,** and Glu/tCr to Gln/tCr was 0.34 in in **TU1.5**, and 0.41 in **PT1.5**. This implies that Glu and Gln ratio hotspot overlap is driven by tNAA decreases.

**Figure 5:**
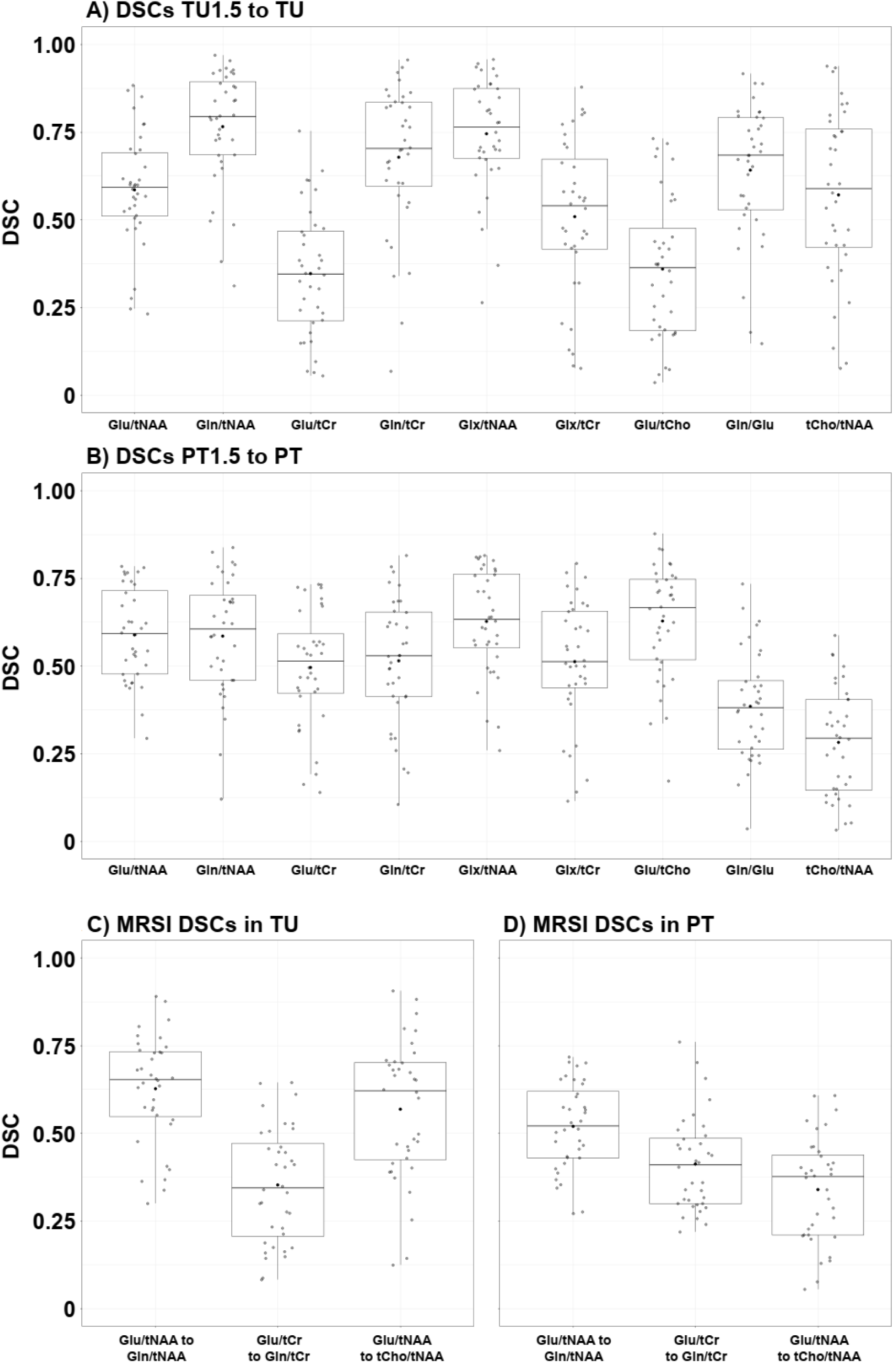
Box plots of all calculated DSCs. DSC = Dice similarity coefficient; Glu = glutamate; Gln = glutamine; Glx = glutamate + glutamine; NAWM = normal-appearing white matter; PT = peritumoral VOI; PT1.5 = peritumoral VOI with 1.5×NAWM threshold; tCho = total choline; tCr = total creatine; tNAA = total N-acetyl aspartate; TU = tumor VOI; TU1.5 = tumor VOI with 1.5×NAWM threshold; VOI = volume of interest.

### Correlations

Of all investigated correlations (Sup.Tbl.6), only eight were over the threshold. Age correlated negatively with IDH-status (-0.70) and positively with Glu/tCr medians in **TU0** (0.57) and Glx/tCr medians in **TU0** (0.54, significant difference discussed above). IDH status correlated negatively with Glx/tCr medians in **TU0** (-0.53). Low or high tumor grade correlated positively with the DSC of Glu/tCr in **TU1.5** (0.57), the Glx/tNAA median in **TU1.5** (0.50), the Glx/tCr median in **TU0** (0.53), and the Gln/tCr median in **TU0** (0.58).

## Discussion

Investigating the peritumoral distribution of Glu and Gln for the first time with high-resolution 7T MRSI, we discovered promising leads into the metabolic imaging of gliomas. Glu and Gln distributions show relevant differences between the compartments while the standard tCho/tNAA cannot discriminate **PT0** and **TU0**, but our interest lies in where Glu and Gln maps diverge. Comparing Glu/tNAA to Gln/tNAA showed that both were significantly different between **PT0**/**TU0/NAWM**, but for Glu/tNAA **PT0** is closer to **TU0** than to **NAWM**. Comparing Glu/tCr to Gln/tCr showed significantly higher values in **PT0** over the other regions in Glu/tCr, while Gln/tCr decreased from **TU0** to **NAWM**. This demonstrated the impact of the chosen reference for metabolic ratios. While Glu/tCr was higher in **PT0**, Glu/tNAA was higher in **TU0**, indicating that the tNAA decrease in the solid tumor masked Glu distribution. Glu/tCho offers an interesting alternative to investigate peritumoral activity as we found it significantly different between **PT0** and **TU0** or **NAWM**, likely due to the contrast of infiltrative interaction with the brain parenchyma versus membrane activity in the solid tumor. Gln/Glu, however, was detectably increased only in **TU0**, which completes the picture that Glu and Gln increases in **PT0** are of similar magnitude, but Gln increases in **TU0** are much more pronounced while Glu relatively decreases.

The most interesting finding for the DSCs was that Glu/tCr and Glu/tCho were higher in **PT1.5** than in **TU1.5**, again showing that separating Glu and Gln by 7T MRSI resolves onco-metabolic heterogeneities. The limited correlations found hint at general increases of Gln and Glu with rising grades that could represent accelerating oncometabolism. The significant difference of Glu/Gln between patients with and without TAE showed the importance of 7T spectral resolution and the potential to investigate tumor/brain interaction.

A final insight is that the application of a hotspot threshold is less necessary than expected to detect differences. Yet a more thorough analysis of glioma compartments and larger sub-cohorts could clarify findings of significance.

Our results agree with previous literature about the role of Glu and Gln in glioma metabolism^5,12,13^. Compared to 3T MRS studies, we confirmed increased tumoral Gln/tCr^21^ but did not find decreased Glu/tCr compared to NAWM^21^. Our findings that high grades correlated with intratumoral Glx/tNAA and Glx/tCr correspond to increasing Glx concentrations between grades 2-4 of the WHO 2017 classification^19^. Our finding of tumoral Glu/tCr positively correlating with increasing grades compares well to a significant correlation of Glu/tCr to survival and progression free survival^22^. We could not replicate the correlation of increasing Glu/tCr-levels^23,24^ with TAE but found peritumoral Gln/Glu to be significant.

Compared to our own previous qualitative research^26^, we could quantize our earlier observations about Glu and Gln increases in gliomas. Two other 7T MRS studies^34,35^ found decreased Glu/tCr and no significant changes for Gln/tCr when comparing tumor to the precuneus but a similar tumoral increase in Gln/Glu as in our work^34^, as well as increased Gln/tCr but decreased Glu/tCr between tumor and NAWM^35^. In total, the state of the art remains unclear due to different MRS methodologies, field strengths, and reference regions.

## Limitations

Our analysis is limited by the cohort size (an issue of nearly all glioma MRS studies) as well as lacking methodological consensus on the definitions of tumor VOIs (such as edemas/infiltration), control regions, hotspots, and gold standards. Relying on metabolic ratios introduces unknowns and can hide/create distinct changes, e.g., NAA decreasing to below the detectability-threshold.

### Outlook

This study showed that 7T MRSI, due to higher resolution and Glu-Gln separation, could investigate peritumoral glioma regions, infiltration, and TAE. In a pathology with abysmal outcomes and the inability of clinical standard MRI to define infiltration extent, we could add value to the clinical description and research into gliomas. For instance we propose the scouting of points of interest for surgical sampling during resection or the monitoring of prospective new drugs such as IDH inhibitors (e.g., Vorasidenib) or glutamate synthesis inhibitors (e.g., Troriluzole). Further ahead, Glu and Gln maps, together with other data such as perfusion maps, could be inputs for synthetic infiltration extent images. This would not be feasible with SVS or low-resolution MRSI.

Methodologically, further developments to investigate the importance of Glu and Gln imaging could include quantitative MRSI, which would require proton density scans as a reference but would allow to disentangle ratios. Continuing to increase our patient cohort and to better define regions for analysis will clarify our findings. Meanwhile, Gln/Glu and Glu/tCho could be explored for their use in the definition of the properties of infiltrative gliomas.

## Supplementary Information

To increase reproducibility, Supplementary Tables 2-5 contain patient-wise medians, VOI volumes, DSCs, and NAWM-based thresholds for the **TU1.5**, **PT1.5**, **TU0**, and **PT0** VOIs.

## Data Availability

Image data produced in the present study are available upon reasonable request to the authors after institutional data clearing house approval. Derived numerical values are included in the supplementary data.

