## Supplements for "Imaging of increased peritumoral glutamate and glutamine in gliomas using 7T MRSI"

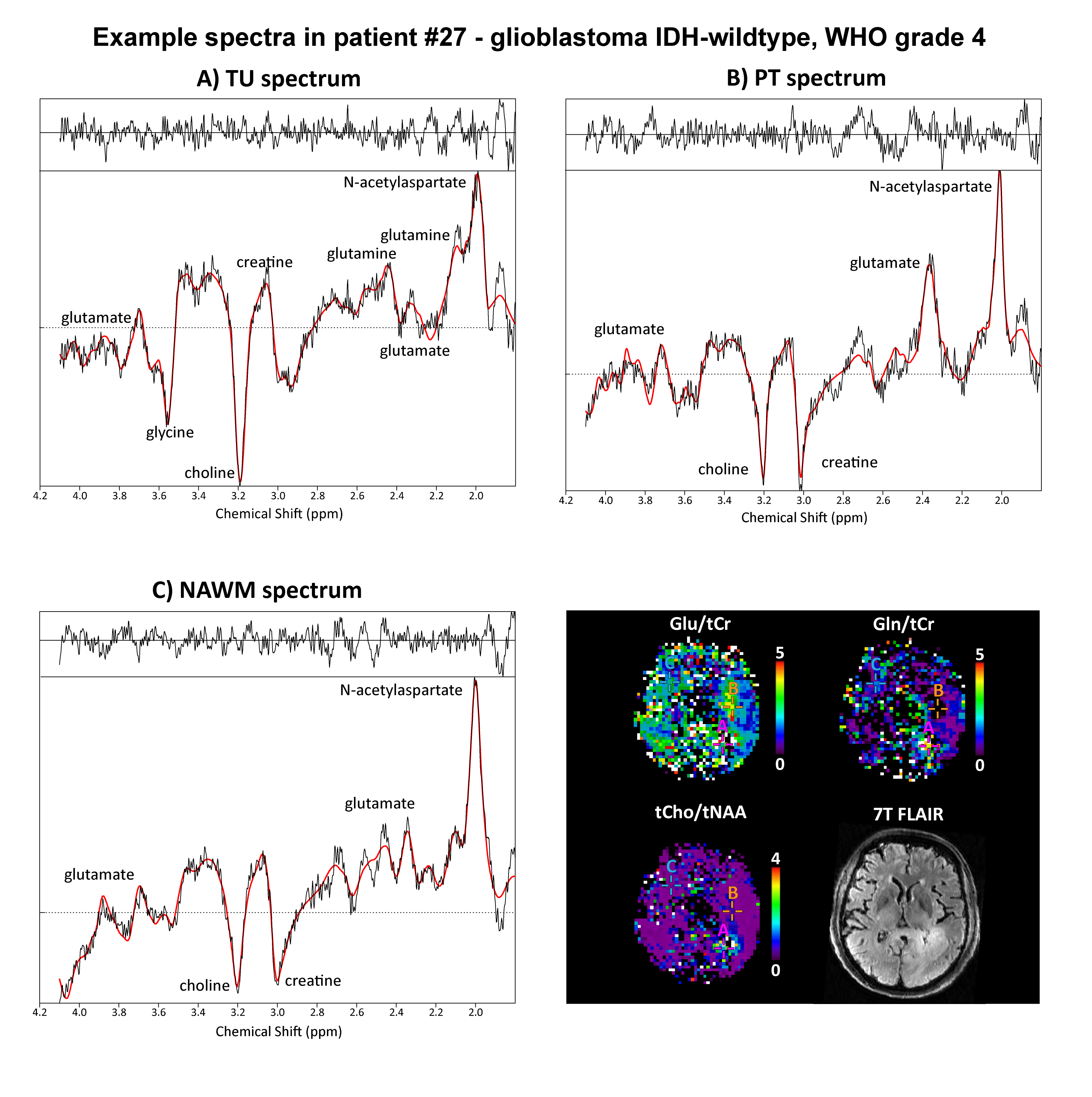

Supplementary figure 1: Exemplary spectra in patient #27 for tumoral, peritumoral and NAWM VOIs. Differences in tCho, Gln, and Glu as well as tNAA are clearly visible. FLAIR = fluid attenuated inversion recovery; Glu = glutamate; Gln = glutamine; IDH = isocitrate dehydrogenase; NAWM = normal-appearing white matter; ppm = parts per million; PT = peritumor; tCho = total choline; tNAA = total N-acetyl aspartate; TU = tumor;

<Table enclosed in tables.xlsx>

Supplementary table 1: MRSinMRS checklist for reproducibility of MRS methods. BW = bandwidth; CH = channel; FID = free induction decay; HFMRC = High Field MR Center; NAWM = normal-appearing white matter; MRS = magnetic resonance spectroscopy; MRSI = magnetic resonance spectroscopic imaging; SNR = signal-to-noise ratio; WET = water suppression enhanced through T1 effects.

<Tables enclosed in tables.xlsx>

Supplementary tables 2,3,4,5: Patient-wise data for all applied thresholds (**TU1.5**, **PT1.5**, **TU0**, **PT0** + **NAWM**) and their medians and IQRs. Presented are DSCs, volumes, ratio medians, and thresholds. Of note, MRSI ratio DSCs in **PT0** and **TU0** are slightly below 1, as the applied spectral quality filtering has removed voxels from the analysis. DSC = dice similarity coefficient; f = female; Glu = glutamate; Gln = glutamine; Glx = glutamate + glutamine; IDH = isocitrate dehydrogenase; IQR = inter-quartile range; NAWM = normal-appearing white matter; m = male; MRSI = magnetic resonance spectroscopic imaging; PT0 = peritumor segmentation without ratio threshold; PT1.5 = peritumor segmentation with 1.5×NAWM threshold; tCho = total choline; tCr = total creatine; tNAA = total N-acetyl aspartate; TU0 = tumor segmentation without ratio threshold; TU1.5 = tumor segmentation with 1.5×NAWM threshold; VOI = volume of interest.

<Table enclosed in tables.xlsx>

Supplementary table 6: Correlations between MRSI ratios and clinical parameters (grade, low grade vs grade, IDH1 mutation, sex, tumor-associated epilepsy, peritumoral seizure onset) for the **TU1.5**, **PT1.5**, **TU0**, and **PT0** VOIs. Glu = glutamate; Gln = glutamine; Glx = glutamate + glutamine; IDH = isocitrate dehydrogenase; NAWM = normal-appearing white matter; MRSI = magnetic resonance spectroscopic imaging; PT0 = peritumor segmentation without ratio threshold; PT1.5 = peritumor segmentation with 1.5×NAWM threshold; tCho = total choline; tCr = total creatine; tNAA = total N-acetyl aspartate; TU0 = tumor segmentation without ratio threshold; TU1.5 = tumor segmentation with 1.5×NAWM threshold; VOI = volume of interest.
